# Seroprevalence of SARS-CoV-2 Antibodies Among Rural Healthcare Workers

**DOI:** 10.1101/2021.06.07.21258375

**Authors:** Jordan Z. Neises, Md Saddam Hossain, Rifat Sultana, Kevin N. Wanniarachchi, Jared W. Wollman, Eric Nelson, Bonny L. Specker, Adam D. Hoppe, Steven R. Lawson, Natalie W. Thiex

## Abstract

The objective of this longitudinal cohort study was to determine the seroprevalence of antibodies to severe acute respiratory syndrome coronavirus-2 (SARS-CoV-2) in healthcare workers employed at healthcare clinics in three rural counties in eastern South Dakota and western Minnesota from May 13, 2020 through December 22, 2020. Three blood draws were performed at five clinical sites and tested for the presence of antibodies against the SARS-CoV-2 virus. Serum samples were tested for the presence of antibodies using a fluorescent microsphere immunoassay (FMIA), neutralization of SARS-CoV-2 Spike-pseudotyped particles (SARS-CoV-2pp) assay, and serum virus neutralization (SVN) assay. The seroprevalence was determined to be 1/336 (0.29%) for samples collected from 5/13/20-7/13/20, 5/260 (1.92%) for samples collected from 8/13/20-9/25/20, and 35/235 (14.89%) for samples collected from 10/16/20-12/22/20. Eight of the 35 (22.8%) seropositive individuals identified in the final draw did not report a previous diagnosis with COVID-19. There was a high correlation (>90%) among the FMIA and virus neutralization assays. Each clinical site’s seroprevalence was higher than the cumulative incidence for the general public in each respective county as reported by state public health agencies. As of December 2020, there was a high percentage (85%) of seronegative individuals in the study population.

## INTRODUCTION

In December of 2019, a cluster of patients in the Wuhan province of China were diagnosed with pneumonia of an unidentified cause. The pneumonia-like disease was caused by a novel beta-type coronavirus, SARS-CoV-2. The newly identified species belongs to the family *Coronaviridae* of the genus *Betacoronavirus*^1^. The disease state caused by SARS-CoV-2 was later renamed COVID-19^2^. SARS-CoV-2 proved difficult to contain due to its high transmissibility, spreading from person to person primarily through respiratory droplets from infected individuals^3,4^. The first reported case of COVID-19 in the United States occurred on January 19, 2020 in Washington state^5^. On March 11, 2020, the World Health Organization (WHO) declared COVID-19 a global pandemic, citing it as the first pandemic caused by a coronavirus^6^. COVID-19 quickly spread throughout highly populated urban areas in the United States. Over 200,000 new cases of COVID-19 were reported in the New York City area within a three-month period^7^.

Early in the pandemic, despite the toll COVID-19 had on urban populations, it remained difficult to predict how the virus would impact rural areas. Notably, seasonal influenza has a shorter duration but higher degree of intensity in rural populations, and regional epidemics frequently begin in urban areas before spreading to rural communities^8^. Furthermore, rural communities often lack the medical resources and infrastructure necessary to handle large-scale outbreaks^9^.

The need for serological testing is required to quantify the true percentage of individuals who were infected with SARS-CoV-2 in the United States. COVID-19 causes a wide range of respiratory symptoms with varying degrees of severity. The elderly and those with preexisting conditions are at higher risk for experiencing a severe case of COVID-19^10,11^. This can be especially worrisome for rural communities where an estimated 17.5% of the population is age 65 or older in comparison with 13.5% in urban communities^12^. Younger individuals without preexisting conditions may present with mild to no symptoms. Unlike symptomatic individuals who often seek medical attention, an asymptomatic individual may not be tested and therefore missed via conventional PCR-based testing protocols. Antibody testing allows for the identification of these asymptomatic and/or undiagnosed individuals.

Healthcare workers are a high-risk group for contracting COVID-19 due to their frequent exposures to infected individuals. In addition, healthcare workers have on-site access to phlebotomy services at the place of work. Therefore, individuals working in the healthcare facilities in eastern South Dakota and western Minnesota were tested for the presence of antibodies against SARS-CoV-2 over the course of an eight-month period.

## METHODS

### Study Design

A longitudinal cohort study was conducted from May 13, 2020 through December 22, 2020 with healthcare workers in eastern South Dakota and western Minnesota. Following recruitment via email, potential participants were guided through an online informed consent process and completed an online questionnaire. Individuals were invited from five clinical sites and were asked to provide three blood samples throughout the course of the study. Participants’ serum was tested for antibodies against SARS-CoV-2. Due to the COVID-19 pandemic, provisions were made in order to ensure the safety of participants and researchers. Each blood draw was performed by clinical staff phlebotomists at the discretion of the clinical site. After collection, the serum samples were turned over to study personnel and tested for SARS-CoV-2 antibodies.

### Participants

Human subjects’ procedures were approved by the South Dakota State University Institutional Review Board prior to the start of the study, were in compliance with relevant laws and institutional guidelines, and were in accordance with the ethical standards of the Declaration of Helsinki. All participants provided informed consent prior to participating in the study.

### Study Sites

Five clinical sites from three different counties were studied in South Dakota and Minnesota. Three clinical sites provide in-patient services, while two are out-patient services only. All three counties are considered non-metro or rural areas based on United States Department of Agriculture (USDA) classification. Two counties have a 2013 Rural-Urban Continuum Code of 9, which indicates completely rural and not surrounded by an adjacent metro area. The third county has a 2013 Rural-Urban Continuum Code of 5 meaning that the population is greater than 20,000 and not surrounded by adjacent metro area^13^.

### Online Questionnaire

Participants provided demographic information including age, sex, race, height and weight via an online questionnaire using QuestionPro (HIPPA certified; Austin, TX). COVID-19 related questions also were asked, including whether they had been previously diagnosed with COVID-19, the date of diagnosis and whether they had exposure to a known positive or suspected positive case of COVID-19.

### Serum collection

Upon collection, participant blood samples were deidentified and incubated at room temperature for 30 min to allow for clotting to occur. Samples were then centrifuged for 10 min at 1,500 x g. Serum was separated from whole blood and stored at -40°C prior to being assayed.

### Fluorescent microsphere immune assay (FMIA)

A quantitative assessment of serum IgG, IgA and IgM antibodies against the SARS-CoV-2 nucleocapsid protein (NCP) was performed using a fluorescent microsphere immunoassay (FMIA) testing platform. To prepare the microspheres for antibody capture, a two-step carbodiimide coupling procedure was used to couple prokaryotic expressed and purified SARS-CoV-2-NCP antigen to Luminex™ microspheres as previously described^14,15^. Briefly, the full length (1257 bp) coding sequence corresponding to the Wuhan-Hu-1, nucleoprotein gene (Genbank MN908947.3) was chemically synthesized and cloned into the bacterial expression plasmid pET-28a (EMD Millipore/Novagen, Billerica, MA). DNA sequencing was used to confirm the identity and in-frame cloning of the SARS-CoV-2 protein with a 6x histidine fusion tag. The recombinant protein was expressed in *Escherichia coli* BL-21 cells and purified using nickel-NTA agarose resin (Qiagen, Valencia, CA). Using a series of titrations, the optimal coupling ratio was calculated to be 25 μg NCP antigen/3.1 × 10^6^ microspheres.

For the performance of the FMIA, 50 μL of heat inactivated serum (diluted 1:50 in PBS-BN) was added to 2.5 × 10^3^ antigen-coupled microspheres. Serum binding IgG, IgA and IgM antibody isotypes were detected using a polyisotypic, anti-human, biotinylated secondary antibody (Invitrogen, Carlsbad, CA) followed by a fluorescent (streptavidin-phycoerythrin) reporter (Invitrogen, Carlsbad, CA) that was added to sample and control wells. Anti-NCP antibodies were quantified through a dual-laser instrument (Bio-Rad Bio-Plex 200) as previously described^16^. The median fluorescent intensity (MFI) for 100 microspheres corresponding to each analyte was recorded for each well, their measurements were mathematically normalized against a serological reference standard to calculate a relative sample-to-positive (S/P) ratio. Determination of a diagnostic sensitivity and specificity threshold cutoff was calculated using a change-point analysis method determined by calculating the mean plus three standard deviations of the negative control, S/P ratios as described^17^. For serum samples tested more than once the mean S/P ratio was calculated for diagnostic determination.

### Serum virus neutralization assay (SVN)

A fluorescent serum virus neutralization assay using live virus was developed for the quantification of neutralizing antibodies produced in response to SARS-CoV-2 infection. Two-fold serial dilutions of heat inactivated serum (1:2 to 1:256) were prepared using MEM + 10% FBS (R&D Systems, Flower Branch, GA) and incubated with an equal volume of SARS-CoV-2 virus strain SDLEMN-20 (South Dakota 2020 isolate) having a titer between 300-400 foci forming units/well and having a final assay range of 1:4 to 1:512. After a 1-h incubation, trypsinized Vero 76 cells were added to the 96-well dilution plate, then incubated at 37°C for 48 h. After incubation, cells were acetone fixed, and virus infected cells were visualized and quantified by staining infected cells with a SARS-CoV-2 nucleoprotein-specific, FITC-conjugated, monoclonal antibody (SD83-108) as described previously^18,19^. Lastly, SVNs were read under a fluorescence microscope and neutralizing antibody titers expressed as the reciprocal of the highest dilution of serum capable of a 90% reduction in florescent foci relative to controls. Both negative and positive control sera were included in all assays.

### Neutralization assay of SARS-CoV-2 Spike-pseudotyped particles (SARS-CoV-2pp)

To mimic the infection condition of human cells, 293T cells were generated, which stably express human ACE2 by lentiviral transduction with pLENTI-hACE2-HygR (a gift from Raffaele De Francesco; Addgene plasmid# 155296). Transduced cells were sorted by flow cytometry 72 h post-transduction based on ACE2 expression detected with anti-hACE2 Alexa fluor 488 conjugated antibodies (Catalog# Fab9332G, clone# 535919, R&D Systems, Minneapolis, MN). After sorting, a population was generated in which 99.3% of the cells expressed ACE2 compared to the parental 293T cells which had no detectable ACE2 expression.

For the purpose of Spike pseudovirus production, the vector pCMV14-3X-Flag-SARS-CoV-2 S was used, a gift from Zhaohui Qian lab (Addgene plasmid #145780)^20^ that carries a codon-optimized cDNA that encodes SARS-CoV2-S glycoprotein (Wuhan 2019) with C-terminal 19 amino acid deletion. Site directed mutagenesis was performed, confirmed by sequencing to make the D614G mutation in spike and named it pCMV-SD614G. Spike pseudovirus particles containing a Luciferase reporter gene were produced in 293T cells by co-transfection of packaging plasmid psAPX2 which was a gift from Didier Trono (Addgene plasmid# 12260), transfer plasmid pLenti-CMV V5-LUC (W567-1) which was gift from Eric Campeau (Addgene plasmid# 21474)^21^ and envelop plasmid pCMV-SD614G in 293T cells using TransIT-Lenti transfection reagent (Mirus Bio, Madison, WI). Plasmids were used at a ratio of 5:3:3 (pspAX2:LUC-blast:SD614G) and a total of 5.5 μg plasmid DNA was mixed in 1 mL of serum free OPTI-MEM media (Gibco, Life Technologies corporation, Grand Island, NY) with 30 μL of transfection reagent. After a 10-min incubation with the transfection reagent, the plasmid mixture was added drop wise onto 293T cells in 100 mm tissue culture dish (Thermo Fisher Scientific, Waltham, MA). After 48 h post transfection, the supernatant containing the released pseudovirus particle was harvested and stored at - 80°C for future use or was used immediately with the assay.

The in-vitro serum neutralization assay was performed as previously described^22^. Briefly, 3.5* 10^3^ ACE2 293T cells were plated on white 96-well cell culture treated plates (Falcon, Waltham, MA) in DMEM supplemented with 10% FBS one day prior to infection with the pseudovirus. To determine the neutralization efficiency, the human sera was serially diluted from 1:20 with a dilution factor of 2 to 8 dilutions in DMEM medium. A volume of 50 μL of spike pseudovirus was added to 50 μL of serum solution and incubated for 1 h at 37° C before adding to the target cells. After 48 h post-infection with the spike pseudovirus, Luciferin (D-Luciferin potassium salt, Goldbio, St. Louis, MO) was added at a final concentration of 200 μg/mL to the cells. Luminescence was measured for each serum dilution using a micro plate reader (BioTekSynergy, Biotech, Winooski, VT). For each well, the luminescence for the highest dilution (1:2560) was divided by 2 to determine a 50% cutoff point. The highest serum dilution that resulted in a 50% neutralization titer (NT_50_) was recorded for the samples.

### Public records search for cumulative incidence data

The cumulative case percentage for each county was accessed via the South Dakota Department of Health and Minnesota Department of Health online database^23,24^. The cumulative case percentage was found for specific dates relevant to individual draw dates at each test site. Public records for county data were accessed on April 1, 2021.

## RESULTS

### Demographic data

In total, 336 healthcare workers from five clinical sites in eastern South Dakota and western Minnesota participated in the study. The majority of study participants were less than 50 years old (69.5%), were primarily women (86.6%), and were of Caucasian race (96.4%; Table 1).

**Table 1.** Demographic characteristics of study participants.

|  | no. (%) |
| --- | --- |
| Age (years) |  |
| 18-29 | 57 (16.9%) |
| 30-39 | 101 (30.0%) |
| 40-49 | 76 (22.6%) |
| 50-59 | 65 (19.3%) |
| 60+ | 37 (11.0%) |
| Gender |  |
| Male | 45 (13.3%) |
| Female | 291 (86.6%) |
| Ethnicity |  |
| Hispanic | 4 (1.2%) |
| Non-Hispanic | 332 (98.8%) |
| Race |  |
| Caucasian or white | 324 (96.4%) |
| Black or African American | 0 (0.0%) |
| American Indian or Alaska Native | 4 (1.2%) |
| Asian | 2 (0.6%) |
| Multiracial | 2 (0.6%) |
| Other | 4 (1.2%) |
| Total participants | 336 |

### Serial blood draws show increasing seropositivity among study participants

Phase 1 blood draws occurred between 5/13/20-7/13/20, and 1/336 (0.3%) healthcare workers were found to have antibodies against SARS-CoV-2 (Table 2). Phase 2 blood draws occurred between 8-13/20-9/25/20, and 5/260 (1.9%) healthcare workers were found to be seropositive (Table 2). All of the individuals who tested positive during Phase 2 had tested negative during Phase 1. Phase 3 blood draws occurred between 10/16/20-12/22/20, and 35/235 (14.8%) healthcare workers were seropositive (Table 2). Four of the five individuals who tested positive in Phase 2 continued with the study and also tested positive during Phase 3.

**Table 2.** A longitudinal estimate of seroprevalence beginning May 13, 2020 through December 22, 2020 reported as the number of positive individuals over the total number tested.

|  | Phase 1 | Phase 2 | Phase 3 |
| --- | --- | --- | --- |
| Draw Sites | (05/13/20 - 07/13/20) | (8/13/20 - 09/25/20) | (10/16/20 - 12/22/20) |
| Site 1 | 1/162 | 3/135 | 7/120 |
| Site 2 | 0/50 | 1/37 | 7/39 |
| Site 3 | 0/13 | - | 4/9 |
| Site 4 | 0/40 | 1/34 | 4/25 |
| Site 5 | 0/71 | 0/54 | 13/42 |
| **Total by phase** | 1/336 (0.3%) | 5/260 (1.9%) | 35/235 (14.8%) |

### Viral neutralization assays confirm FMIA results and show study participants have neutralizing antibodies

The FMIA identified 32 positive samples from Phase 3. These 32 samples along with 3 borderline positive serum samples, and a subset of 18 negative samples were subjected to confirmatory testing via SARS-CoV-2 Spike-pseudotyped particles assay (SARS-CoV-2pp) and serum virus neutralization assay (SVN), two assays designed to identify neutralizing antibodies specific to SARS-CoV-2. Neutralizing antibodies were identified in 35 samples using SVN and 32 samples using SARS-CoV-2pp assays. The average positive titer for SVN and SARS-CoV-2pp was 1:81 and 1:191, respectively (Supplemental Table 1). Samples were deemed “positive” if they were positive for two out of the three assays (FMIA, SVN, SARS-CoV-2pp). In total, 53 samples were tested with 91.4% agreement between each of the neutralizing assays and between the neutralizing assays and the FMIA (Table 3, Supplemental Table 1). *Antibody testing shows higher seroprevalence than county-level cumulative incidence reported by state health departments*

**Table 3.** Phase 3 assay comparison.

|  |  | FMIA |  |  | SARS-CoV-2pp |  |  |
| --- | --- | --- | --- | --- | --- | --- | --- |
|  |  | Positive | Negative | Total | Positive | Negative | Total |
| **SVN** | **Positive** | 32 | 3 | 35 | 32 | 3 | 35 |
|  | **Negative** | 0 | 18 | 18 | 0 | 21 | 21 |
|  | **Total** | 32 | 21 | 53 | 32 | 21 | 53 |
| **SARS-CoV-2pp** | **Positive** | 29 | 3 | 32 |  |  |  |
|  | **Negative** | 3 | 18 | 21 |  |  |  |
|  | **Total** | 32 | 21 | 53 |  |  |  |

FMIA (fluorescent microsphere immunoassay), SVN (serum virus neutralization), SARS-CoV-2pp (SARS-CoV-2 Spike-pseudotyped particles)

The seroprevalence for each clinical site in Phase 3 was compared to the cumulative case rate for their respective county on the date of the blood draw. Each of the clinical sites had a higher seroprevalence amongst their healthcare workers than the cumulative case rate reported by the state department of health on the same date (Table 4).

**Table 4.** Comparison of cumulative cases in the general population and seroprevalence in the study population at the time of the final blood draw.

| **Location** | **Date** | **\*Cumulative cases by County/Date %** | **Clinical Site Seroprevalence % (# positive/total)** |
| --- | --- | --- | --- |
| Site 1 | 10/16/2020 | 3.1% | 5.8% (7/120) |
| Site 2 | 11/03/2020 | 4.5% | 17.9% (7/39) |
| Site 3 | 12/23/2020 | 8.1% | 44.0% (4/9) |
| Site 4 | 12/03/2020 | 7.7% | 16.0% (4/25) |
| Site 5 | 12/03/2020 | 7.0% | 30.9% (13/42) |

\* Public data retrieved from South Dakota Department of Health (https://doh.sd.gov/COVID/Dashboard.aspx); and Minnesota Department of Health (https://www.health.state.mn.us/diseases/coronavirus/index.html)

### Asymptomatic or undiagnosed infection and occupational COVID-19 exposures

In Phase 1, the one participant who was seropositive also indicated having been diagnosed with COVID-19 prior their first blood draw. Of the healthcare workers sampled in Phase 1, 15.1% had contact with patients confirmed of having an active case of COVID-19 (Table 5). In Phase 2, three participants indicated that they had been diagnosed with COVID-19 prior to having their blood drawn. A total of 44.5% study participants had direct contact with patients confirmed of having an active COVID-19 infection (Table 5). In Phase 3, 15.4% indicated having been diagnosed with COVID-19 (Table 5) prior to having their blood drawn and 69.1% reported having direct contact with patients confirmed of having an active case of COVID-19 (Table 5).

**Table 5.** COVID-19 diagnosis and self-reported occupational exposure data

|  | Total |
| --- | --- |
| Questionnaire 1: |  |
| Study participant diagnosed with COVID-19 | 1 (0.29%) |
| Direct contact with patients confirmed of having an active COVID -19 infection | 50 (15.1%) |
| No direct contact with patients suspected *or* confirmed of having COVID-19 | 185 (55.0%) |
| Total participants | 336 |
| Questionnaire 2: |  |
| Study participant diagnosed with COVID -19 | 3 (1.5%) |
| Direct contact with patients confirmed of having an active COVID -19 infection | 88 (44.5%) |
| No direct contact with patients suspected *or* confirmed of having COVID-19 | 75 (38.4%) |
| Total participants | 195 |
| Questionnaire 3: |  |
| Study participant diagnosed with COVID -19 | 27 (15.4%) |
| Direct contact with patients confirmed of having an active COVID -19 infection | 121 (69.1%) |
| No direct contact with patients suspected *or* confirmed of having COVID-19 | 50 (28.5%) |
| Total participants | 175 |

Of the 35 individuals who tested positive for antibodies during Phase 3, 27 (77.1%) reported having been diagnosed with COVID-19 prior to having their blood drawn and 8 (22.8%) indicated they had not been previously diagnosed with COVID-19.

In Phase 3, seventeen of the 121 (14%) study participants who reported having direct contact with COVID-19 patients were seropositive; whereas, 9 of the 50 (18%) study participants who reported having no direct contact with patients suspected or confirmed of having COVID-19 were seropositive. These results indicate that occupational exposure to COVID-19 is not a predominant dynamic for testing positive in this population.

### Participant retention and loss to follow up

The single seropositive individual from Phase 1 was lost to follow-up and did not return for subsequent draws. Overall, 76 participants from the Phase 1 of testing were lost to follow-up (Figure 1). Four of the five individuals who were found to have antibodies in Phase 2 returned for the final blood draw.

**Figure 1.**
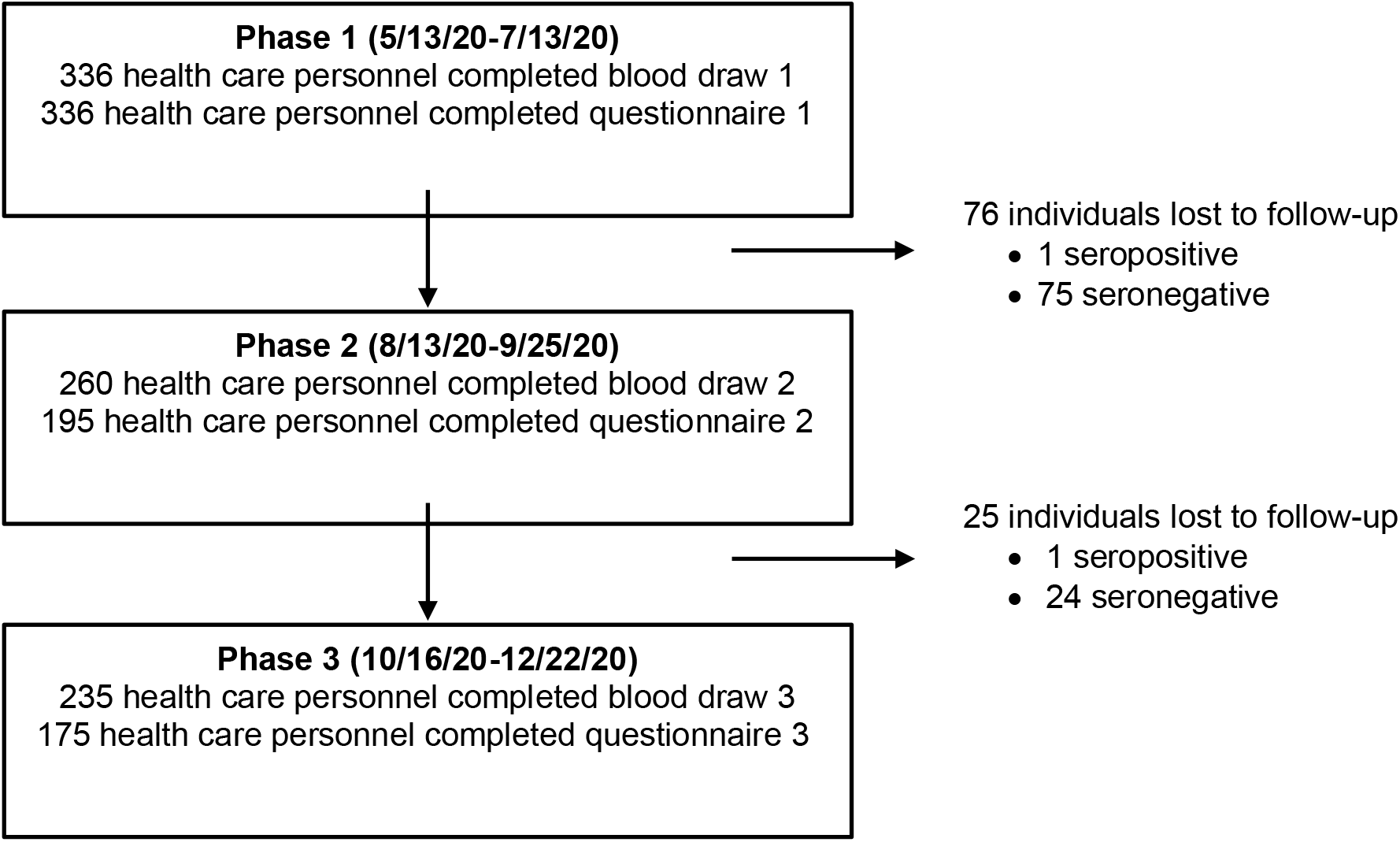
Retention and loss of study participants.

## DISCUSSION

In conclusion, 14.8% of healthcare workers from eastern South Dakota and western Minnesota seroconverted over the course of an 8-month testing period while 85.1% remained susceptible to COVID-19. Of individuals who tested positive for SARS-CoV-2 antibodies, 22.8% had not been diagnosed with COVID-19. Greater than 90% of the seropositive individuals had neutralizing antibodies. Healthcare workers with direct occupational exposure to COVID-19 patients had similar seroprevalence rates compared to healthcare workers with no contact to known or suspected COVID-19 patients (14% vs. 18%, respectively at Phase 3 visit). Healthcare workers enrolled in the study had a higher seroprevalence of SARS-CoV-2 exposure when compared to county-wide cumulative incidence data as reported by the state Departments of Health. However, at the end of the testing period there still remained a large majority of individuals who did not have a detectable antibody response suggesting they were still susceptible to infection. COVID-19 vaccines became available to healthcare workers shortly after the end of the Phase 3 testing period.

Healthcare workers had access to COVID-19 diagnostic testing via their employer, yet 22.8% of seropositive individuals were not previously diagnosed. This percent of undiagnosed individuals is similar to other published reports. A large-scale study involving 109,076 participants in the United Kingdom found that 30.6% of seropositive individuals were not previously diagnosed^25^, and 10.3% of seropositive healthcare workers in the New York City Area also went undiagnosed^26^. The reason these workers went undiagnosed is not known, but could be due to having an asymptomatic infection, were not tested, or had a false negative test result.

There was greater than 90% correlation among the FMIA and two neutralizing antibody assays. The SARS-CoV-2pp assay used a higher starting dilution of 1:20. If a lower starting dilution been used, one would likely see neutralizing activity in all 35 positive samples from Phase 3. In this study, greater than 90% of the seropositive individuals had neutralizing antibodies. The percentage of seropositive individuals displaying neutralizing activity is variable with published reports ranging from 33% to 75%^27,28^.

There was a high level of SARS-CoV-2 viral transmission in the three counties where the study locations resided. All three counties reported substantial community spread (100 cases/100,000 population per week) starting in mid-to late-August and continuing for the duration of the study, with peak cases per week from ∼750/100,000 in one county to over 1,100/100,000 in another county.

There are multiple strengths to this longitudinal study. Serially testing study participants over an eight-month period allowed observation of seroconversion in study participants over time and for comparison to previous negative results within an individual. Using healthcare workers as study participants allowed for safe human subjects research to occur during a global pandemic since staff phlebotomist performed blood draws at each site, thereby ensuring limited face-to-face contact between participants and the research team. Another strength was the decision to include multiple study locations. Two of three counties housing testing sites are considered class 9 rural areas, with populations of less than 2,500. The third county is considered a class 5 rural area with a population of more than 20,000 people (Cromartie, 2020). The overall seroprevalence from the five separate testing sites provides a more reasonable estimate compared to a single site which may be prone to isolated outbreaks.

A limitation of the study was the potential for participant bias as individuals who suspected they were exposed to SARS-CoV-2 may have been more likely to continue the study. This coincides with the fact that most participants reported having direct contact with either a suspected or a confirmed COVID-19 case (71.4%) in Phase 3. This exposure rate is likely higher than that among the general population given the estimated case rates for each county were below 9% at the time of this study. The study began with 336 participants with 101 individuals lost to follow-up prior to the final blood draw. The retention rate for blood draws over the course of the study was 69.9% with 74.7% of those with a final blood draw also completing the final questionnaire.

In summary, serological testing will continue to be an important metric for understanding the true number of individuals who have been exposed to the SARS-CoV-2 virus and will be an essential tool for understanding the scope to which COVID-19 has spread in rural populations.

## Data Availability

Data is available upon request.

## ACKNOWLEDGEMENTS

We thank the clinical staff liaisons and phlebotomists, as well as the participants, for their collecting and offering blood samples for the study.

---

**Supplementary table 1.** Positive sample confirmation comparison among three separate assays.

| **Sample ID** | **Diagnosed with Covid-19** | **Seropositive via FMIA** | **SVN Titer** | **SARS-CoV-2pp Titer** |
| --- | --- | --- | --- | --- |
| Sample 1 | No | Yes | 1:256 | 1:640 |
| Sample 2 | Yes | Yes | 1:256 | 1:640 |
| Sample 3 | Yes | Yes | 1:256 | 1:640 |
| Sample 4 | Yes | Yes | 1:256 | 1:640 |
| Sample 5 | Yes | Yes | 1:64 | 1:640 |
| Sample 6 | Yes | Yes | 1:128 | 1:640 |
| Sample 7 | No | Yes | 1:128 | 1:320 |
| Sample 8 | Yes | Yes | 1:32 | 1:320 |
| Sample 9 | Yes | Yes | 1:128 | 1:160 |
| Sample 10 | Yes | Yes | 1:256 | 1:160 |
| Sample 11 | Yes | Yes | 1:64 | 1:160 |
| Sample 12 | Yes | Yes | 1:128 | 1:160 |
| Sample 13 | Yes | Yes | 1:32 | 1:160 |
| Sample 14 | Yes | Yes | 1:32 | 1:160 |
| Sample 15 | Yes | Yes | 1:32 | 1:160 |
| Sample 16 | Yes | Yes | 1:64 | 1:160 |
| Sample 17 | Yes | Yes | 1:32 | 1:160 |
| Sample 18 | Yes | No | 1:16 | 1:80 |
| Sample 19 | Yes | Yes | 1:128 | 1:80 |
| Sample 20 | Yes | Yes | 1:32 | 1:80 |
| Sample 21 | Yes | Yes | 1:32 | 1:80 |
| Sample 22 | No | Yes | 1:64 | 1:80 |
| Sample 23 | Yes | Yes | 1:32 | 1:80 |
| Sample 24 | Yes | Yes | 1:32 | 1:80 |
| Sample 25 | Yes | Yes | 1:64 | 1:80 |
| Sample 26 | Yes | No | 1:32 | 1:40 |
| Sample 27 | No | Yes | 1:32 | 1:40 |
| Sample 28 | No | Yes | 1:16 | 1:40 |
| Sample 29 | Yes | Yes | 1:32 | 1:40 |
| Sample 30 | Yes | Yes | 1:64 | 0 |
| Sample 31 | No | Yes | 1:16 | 1:40 |
| Sample 32 | Yes | Yes | 1:16 | 0 |
| Sample 33 | Yes | Yes | 1:32 | 0 |
| Sample 34 | No | No | 1:8 | 1:20 |
| Sample 35 | Yes | Yes | 1:64 | 1:20 |
| Sample 36 | No | No | 0 | 0 |
| Sample 37 | No | No | 0 | 0 |
| Sample 38 | No | No | 0 | 0 |
| Sample 39 | No | No | 0 | 0 |
| Sample 40 | No | No | 0 | 0 |
| Sample 41 | No | No | 0 | 0 |
| Sample 42 | No | No | 0 | 0 |
| Sample 43 | No | No | 0 | 0 |
| Sample 44 | No | No | 0 | 0 |
| Sample 45 | No | No | 0 | 0 |
| Sample 46 | No | No | 0 | 0 |
| Sample 47 | No | No | 0 | 0 |
| Sample 48 | No | No | 0 | 0 |
| Sample 49 | No | No | 0 | 0 |
| Sample 50 | No | No | 0 | 0 |
| Sample 52 | No | No | 0 | 0 |
| Sample 52 | No | No | 0 | 0 |
| Sample 53 | No | No | 0 | 0 |

## Notes

### Competing Interest Statement

The authors have declared no competing interest.

### Funding Statement

South Dakota Governor's Office of Economic Development, BioSNTR

### Author Declarations

Human subjects procedures were approved by the South Dakota State University Institutional Review Board

